# Perturbation of the Preterm Human Immune System in Early Life

**DOI:** 10.1101/2025.09.03.25334934

**Authors:** Benjamin A. Fensterheim, Michelle McKeague, Divij Mathew, Shwetank, Ajinkya Pattekar, Sean Nasta, Macy Kee, Cynthia Clendenin, Zachary Martinez, Caroline Diorio, Allison R. Greenplate, Krithika Lingappan, E. John Wherry

**Affiliations:** Department of Pediatrics, Division of Neonatology, Children’s Hospital of Philadelphia, Philadelphia, PA, USA; Department of Systems Pharmacology and Translational Therapeutics, University of Pennsylvania Perelman School of Medicine, Philadelphia, PA, USA; Institute for Immunology and Immune Health, University of Pennsylvania Perelman School of Medicine, Philadelphia, PA, USA; Department of Pediatrics, Division of Oncology, Children’s Hospital of Philadelphia, Philadelphia, PA, USA

**Author notes:** Corresponding Authors: E. John Wherry, Ph.D. University of Pennsylvania 421 Curie Blvd, Philadelphia, PA 19104, Benjamin A. Fensterheim, M.D., Ph.D. Children’s Hospital of Philadelphia 3401 Civic Center Blvd, Philadelphia, PA 19146.

## Abstract

Although inflammatory complications are common in preterm infants, their effects on neonatal immune development remain poorly defined. We therefore investigated whether severe bronchopulmonary dysplasia (BPD) and systemic infection, two major complications of prematurity, produce distinct immune signatures and change immune composition over time. We performed longitudinal high-dimensional immune profiling of residual whole blood from 38 preterm infants sampled every two weeks, along with 10 term infants at birth. Preterm infants with severe BPD showed a progressive increase in Th17-polarized CD4^+^ T cells, neutrophils, and Th17-related cytokines compared to age-matched infants with moderate BPD. In contrast, some preterm infants with systemic bacterial or viral infections mounted robust CD8^+^, CD4^+^, and γδ T cell responses, with oligoclonal expansion, terminal differentiation, and coordinated plasma cytokine shifts that persisted well beyond resolution of infection. These findings demonstrate that different preterm comorbidities imprint the neonatal immune system in divergent ways. Longitudinal immune profiling offers a powerful tool to uncover these trajectories and identify potential targets for intervention.

## Introduction

During gestation, the human immune system develops along a programmed trajectory that prepares the fetus for birth^1^. This early-life immune system is not simply an immature version of the adult counterpart but rather a dynamic and unique network suited to support organ development and establish homeostasis^2–5^. Early-life immune cells arise from distinct hematopoietic precursors^6,7^ and have unique responses to pro-inflammatory stimuli^8,9^ when compared to adult immune cells. Early-life T cells have a more limited T cell receptor (TCR) repertoire^10^, are slower to generate long-lived memory populations^11,12^, release different cytokines^13^, and polarize in unique patterns compared to adult T cells^14,15^. After term birth, this specialized immune system interfaces with numerous evolutionarily expected stimuli, such as commensal microbes, which ultimately guide the development of the immune system along a stereotypic path towards adult maturity^16,17^.

Preterm birth disrupts this process of immune development, exposing infants to both natural and iatrogenic stressors before immune and organ systems are prepared for extrauterine life^18^. These stressors, often required for survival in the neonatal intensive care unit (NICU), include but are not limited to hyperoxia, positive pressure mechanical ventilation, commensal and pathogenic microbes, broad-spectrum antibiotics, and parenteral nutrition^19–23^. Ultimately, these exposures can predispose to developmental diseases around the body such as bronchopulmonary dysplasia (BPD) in the lungs, necrotizing enterocolitis (NEC) in the gut, or systemic bacterial and viral infections, conditions that themselves often require additional supportive yet inflammatory interventions^24^. How the rapidly evolving preterm immune system responds to such stressors is poorly defined and often extrapolated from work on term infants, yet there is evidence that such stressors may alter the developmental trajectory of the early-life immune system. Microbial dysbiosis, which occurs more commonly in infants with clinical illness, may change otherwise stereotypic immune development^16,25^. Infants who suffer from clinical illness in early life have an altered risk of allergic, autoimmune, and infectious diseases later in life^26–30^. Despite these observations, how preterm birth and its complications durably alter the developmental trajectory of the immune system remains unclear.

Here, we aimed to define the developmental trajectory of the preterm immune system with a focus on interrogating how different comorbidities of prematurity might alter this trajectory. We leveraged residual whole blood derived from complete blood counts (CBC) sent from infants in the NICU and performed serial immune profiling of cells and proteins on 38 preterm infants every two weeks, as well as 10 term infants at birth. This granular approach identified immune features that changed over time with gestation, revealing patterns of immune components, cell activation and dynamics, and immune network changes that reflect gestational trajectory and others that diverge due to preterm postnatal life. Moreover, these analyses identified profound immune perturbations associated with preterm comorbidities, such as infection and severe BPD, and highlighted distinct and long-lasting developmental immune changes. These findings illuminate the interplay between immune ontogeny and disease in preterm infants, highlight potential directions for new interventions to improve outcomes, and demonstrate the value of frequent high-dimensional immune profiling in capturing novel and diverse features of preterm disease.

## Results

### Residual blood from preterm infant clinical samples yields high-fidelity, high-dimensional immune information

To begin to interrogate preterm immune system development, we first established a pipeline to capture residual whole blood associated with routine clinical assessments of hospitalized preterm infants at the Hospital of the University of Pennsylvania (HUP) Intensive Care Nursery (ICN), blood that would have otherwise been discarded (**Figure 1A**). In the HUP ICN, it is common clinical practice to send whole blood for CBC assessment approximately every two weeks on all hospitalized preterm infants to monitor for anemia of prematurity. This clinical practice provided an opportunity to test whether residual, unused blood could provide material for detailed assessment of the developing immune system in preterm infants. We enrolled 38 preterm infants born between 23 weeks and 0 days and 32 weeks and 6 days gestation; we excluded infants with known immunologic or genetic conditions, or who were born to mothers infected with human immunodeficiency virus (HIV). We also enrolled 10 otherwise healthy term infants who had a single CBC sent for evaluation of hyperbilirubinemia in the newborn nursery at HUP. The infants in the study spanned the gestational age and weight spectrum, and the preterm infants experienced different clinical comorbidities during their time in the HUP ICN (**Figure 1B**). We then obtained residual blood from the clinical laboratory, with the first sample collected for most infants within 48 hours of life. We continued to collect available blood approximately every two weeks, with a minimum of 12 days between samples. We stopped collecting samples once infants reached a corrected gestational age (cGA) of 40 weeks or at discharge if earlier than this age. Infants had different numbers of serial samples collected that were dependent on their total time in the HUP ICN (**Figure 1C**).

**Figure 1:**
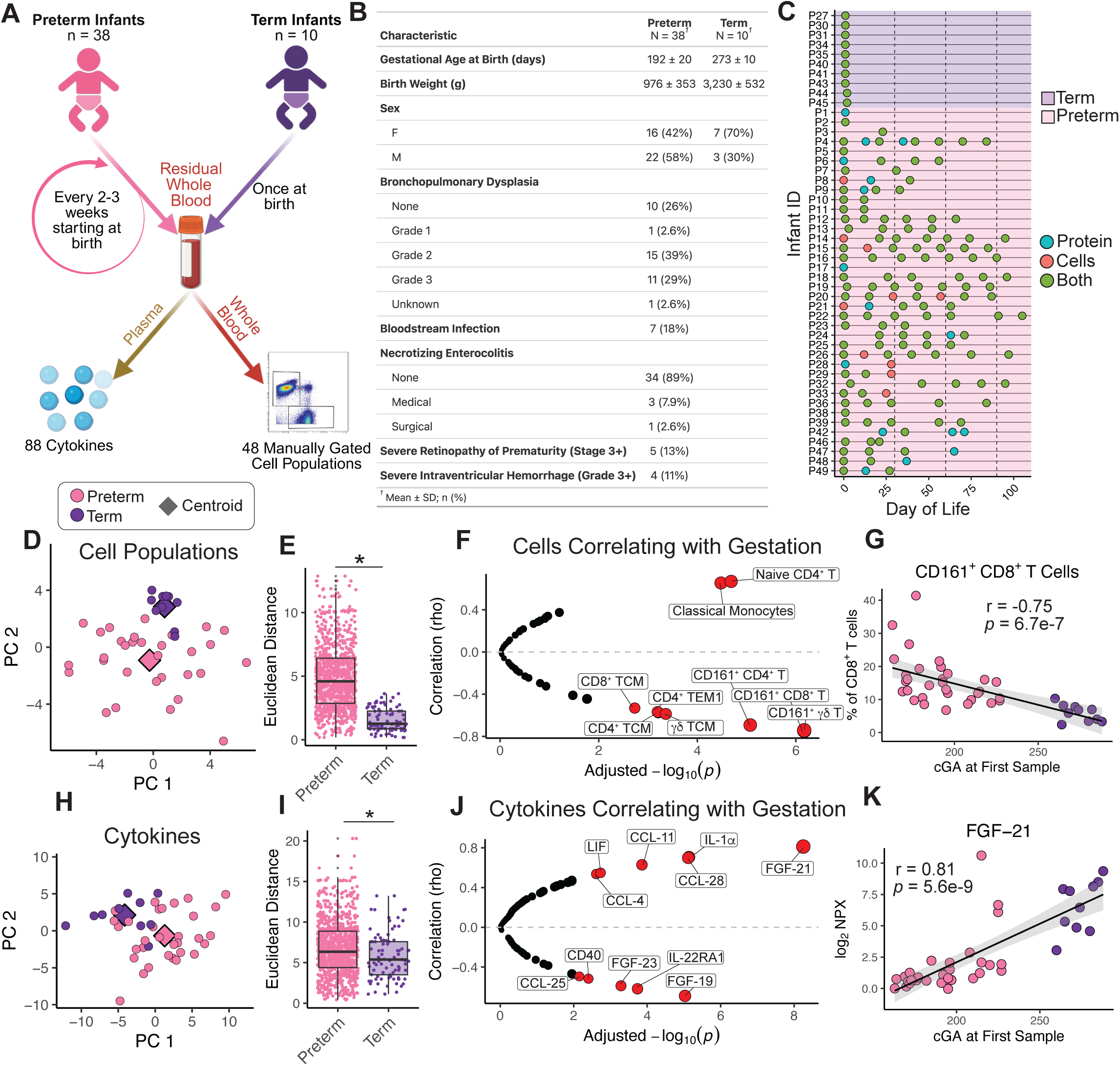
Immune cells and proteins change over gestation. A) Overview of the study design. B) Overview of the clinical features of the infants in the study, including clinical comorbidities during the observation period. C) Each sample as collected from each infant in the study. Each dot represents a single sample, and the color of the dot represents whether the sample was used for Olink only (Protein), CyTOF only (Cells), or both. D) PCA plot of cell populations from the first samples of life for each infant. Dots colored categorically by term or preterm. Purple and pink diamonds represent the centroid of term and preterm groups, respectively. E) For cell populations, Euclidean distances between each PCA coordinate within a group to all other points in that group. F) Plot showing cell populations that significantly correlate with corrected gestational age at birth. Red indicates statistically significant as determined by Benjamini-Hochberg (BH) adjusted p-values < 0.01. G) Linear regression of CD161^+^ CD8^+^ T cells over gestation. Spearman correlation (rho) and p-value shown. H) PCA plot of proteins from the first samples of life for each infant. I) Euclidean distances for cytokines between each PCA coordinate within a group to all other points in that group. L) Plot showing cytokines that significantly correlate with corrected gestational age at birth. Significance determined by BH adjusted p-values < 0.01) Linear regression of FGF-21 over gestational age. Pearson correlation and p-value shown.

Residual blood samples were processed to obtain plasma for proteomic analysis using the Olink platform and for whole blood mass cytometry by time-of-flight (CyTOF) to interrogate circulating leukocyte populations. Each sample provided ∼100-300μL of remaining material after clinical analysis. CyTOF analysis identified 15 major immune cell populations and 33 sub-populations (see gating strategy in **Supplemental Figure 1**). Using plasma from these samples we were able to resolve 88 immune proteins using Olink. Some collected samples did not have sufficient blood volume for any analysis, and some only had sufficient volume for CyTOF or Olink (**Figure 1C**).

Residual blood samples were obtained from the clinical lab within 24-36 hours of collection. Given the potential variability in such residual samples, including possible effects of time since collection on leukocyte populations, we first examined the frequencies and activation patterns of major immune cell types, including sensitive populations like neutrophils and eosinophils. These data indicated that, at least within the collection times of this study, the frequencies of major peripheral blood leukocyte populations were stable and did not display major changes or variability associated with time since collection or volume of residual blood available for analysis (**Supplemental Figure 2A** and **Supplemental Figure 2B**). Thus, we were able to map longitudinal trajectories of 30+ immune cell populations for each preterm infant.

### Immune features at birth vary with gestational age

How the immune cell and protein composition changes throughout gestation has been extensively studied in placental umbilical cord blood^31–33^. Although these data provide some insight into the state of the infant immune system during birth, there is emerging evidence that umbilical cord blood does not reliably reflect the immune composition of infant blood immediately after birth, likely because umbilical cord blood is capturing many changes that occur during the birthing process^16,34^. As a result, there are limited assessments of how gestational programming shapes the composition of infant peripheral blood at birth. To begin to investigate the effect of gestational programming on the infant immune system at birth, we first analyzed samples collected within 0-48 hours of birth, typically the first CBC of life, one from each preterm or term infant. To assess the global landscape of the immune cell composition at birth for each infant, we applied principal component analysis (PCA) for circulating leukocyte populations. The overall immune landscape map of term and preterm infants differed substantially, generating divergent PCA centroids (**Figure 1D**). The interindividual distance between the cell populations of preterm infants was significantly greater than term infants (**Figure 1E**), potentially reflecting the broader gestational age spectrum of the preterm infant population compared to the term infants or more variable indications for delivery.

We next asked what immune features might contribute the most to this intraindividual immune heterogeneity in early life. We first examined how the frequency of individual immune populations correlated with gestational age. CD8^+^, CD4^+^, and ψ8 T cells that express CD161 (KLRB1), as well as non-naïve T cell subsets, steadily decreased in abundance from the earliest to latest gestational ages, whereas the proportion of classical monocytes as well as naïve CD4^+^ T cells as a fraction of total CD4^+^ T cells increased over development (**Figure 1F** and **1G**). CD161 is a marker found on Th17/Tc17 biased T cells and the abundance of CD161^+^ T cells inversely correlates with gestational age in studies of umbilical cord blood^31,35,36^. The identified pattern of these cells here suggests that this correlation continues in the peripheral blood of preterm infants immediately after birth.

To complement the cellular data, we also examined differences in circulating immune proteins in plasma associated with preterm birth and gestational age. Indeed, PCA of plasma cytokines also distinguished term and preterm infants (**Figure 1H**). Again, the interindividual distance in PCA space based on plasma proteins was greater for preterm infants compared to term infants (**Figure 1I**). Several cytokines and circulating mediators differed across gestational age (**Figure 1J**), including fibroblast growth factor (FGF) 19 and 21, which help regulate metabolism in the liver, increased and decreased respectively (**Figure 1K**). FGF23, a regulator of bone phosphate metabolism, decreased. There were also changes in chemokines (CCL4, 11, 25, and 28) and pro-inflammatory cytokines like interleukin 1-alpha (IL-1α). These data demonstrate that residual infant blood can effectively capture high-dimensional immune cell and protein composition and highlight how gestational programming continues to imprint major features of the immune system immediately after birth.

### Preterm immune features deviate from gestational development in postnatal life

Although many aspects of solid organ development continue along a programmed developmental trajectory after preterm birth, conditions of postnatal life can redirect this course. These deviations can manifest as clinical disease and can have long-term clinical consequences^24,37^. However, in contrast to solid organs, relatively little is known about how postnatal life impacts the immune system in preterm infants, despite the real possibility that such deviations could also contribute to disease^2,18,38,39^. We therefore next asked which features of the immune system diverge from their gestational programming in response to postnatal life in the NICU. To address this question, we compared two regression models. The first model, calculated in **Figure 1**, is derived from the first sample of life in preterm and term infants and represents the effect of gestational programming on individual immune features. The second model is derived from all postnatal samples collected from preterm infants and represents the effect of postnatal life on these same features. To generate this second model, we built a linear mixed-effects model for each immune feature using all longitudinal samples from each preterm infant. This approach accounted for repeated measures within individuals and incorporated this longitudinal information. We then projected the fixed effects of the model, reflecting change over time since birth independent of individual variation, onto a line. Finally, we compared the scaled slopes from this postnatal line to the line generated from the gestational model. Because both trajectories spanned a similar range of corrected gestational ages, as preterm infants exited the study at term-equivalent age and the gestational model included term infants, this comparison allowed us to assess whether each immune feature remained on its expected gestational course or deviated due to postnatal influences.

The frequency of some immune cell populations significantly deviated from their gestational trajectory, whereas others did not. For example, the proportion of neutrophils, naïve CD8^+^, CD4^+^ and ψ8 T cells steadily decreased over time in most infants (**Figure 2A**). Conversely, the proportion of many sub-populations of non-naïve ψ8 T cells and conventional T cells, as well as conventional DCs and B cells, increased over postnatal time in the NICU. Some cells, such as CD161^+^ CD8^+^ T cells, continued to closely follow a gestational trajectory in postnatal life. This pattern was also found among the plasma cytokines, particularly among proteins related to organ and vascular development such as an increase in the bone-resorption-promoting receptor of activator of NF-kB ligand (RANKL), as well as a decrease in cytokines such as hepatocyte growth factor (HGF) and vascular endothelial growth factor A (VEGF-A) (**Figure 2B**). Many cytokines, such as FGF-21, appeared resistant to deviation from the gestational trajectory in postnatal life.

**Figure 2:**
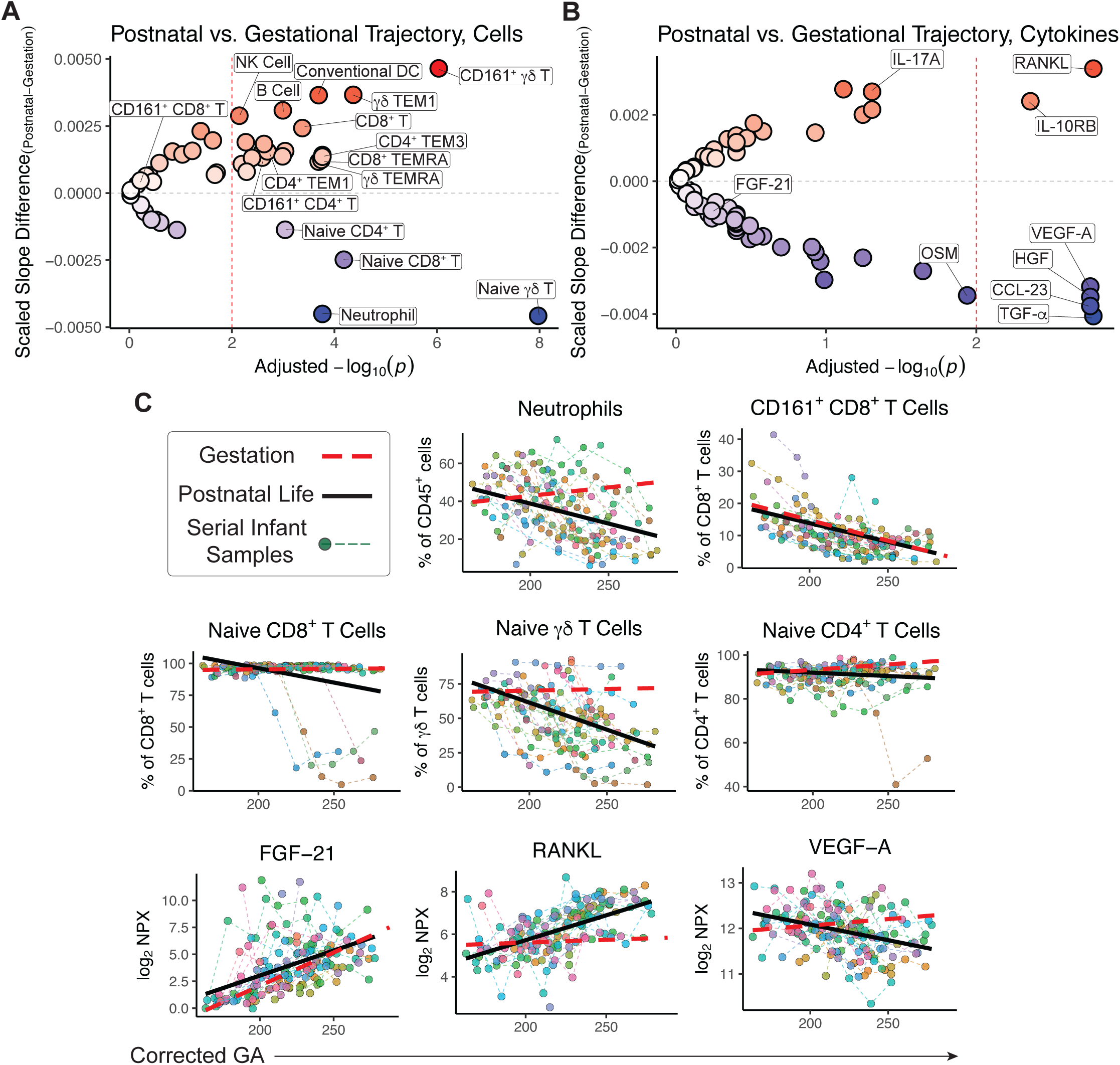
Postnatal life alters the gestational immune trajectory in preterm infants. Linear mixed models were generated for each cell and cytokine. A) The difference of the scaled slopes from the regression generated from all preterm samples and the regression generated from only at-birth samples, for each cell and B) for each cytokine. Cell types above the center dashed line represent populations whose slope increases compared to the expected gestational age progression for that cell type at birth, and cell types below the center dotted line represent populations whose slope decreases. Color of the dots represents the strength of the slope deviation. Significance set at p <0.01 of BH-adjusted p values as determined by the difference in slopes of the two linear models. C) Models of specific cell populations neutrophils, CD161^+^ CD8^+^ T cells, naïve CD8^+^ T cells, naïve γ8 T cells, and naïve CD4^+^ T cells, as well as cytokines FGF-21, RANKL and VEGF-A, across cGA time. For each plot, the solid black line represents the regression derived from all collected preterm samples. The red dashed line represents the regression derived from the at-birth term and preterm samples only, as calculated in Figure 1. Each dot color corresponds to a single preterm infant and the dashed lines connect serial samples collected from the same infant over time.

Immune features deviated from their gestational trajectories with distinct patterns and dynamics. Some changes were observed consistently across the entire preterm cohort over time, whereas others occurred only in a subset of infants (**Figure 2C**). For example, the proportion of neutrophils, naïve ψ8 T cells, and circulating concentrations of VEGF-A were altered from gestational programming in most preterm infants (**Figure 2C**). In contrast, there were other patterns, such as a gradual decrease in the proportion of naïve CD4^+^ T cells, and a sudden and robust drop in the proportion of naïve CD8^+^ T cells, that occurred in a smaller number of preterm infants. These data indicate that postnatal life is associated with major deviations from gestational immune system development, with some changes common to most or all preterm infants and others only in a subset of preterm infants, suggesting that specific early-life postnatal conditions may drive immune responses in some infants.

### Severe bronchopulmonary dysplasia is associated with divergent Th17 activity

We next asked whether diagnosed comorbidities of prematurity were associated with some of the deviations of the immune system in postnatal life. The most prevalent preterm comorbidity in the cohort was, expectedly, BPD. BPD is a respiratory disease of prematurity that is defined by distorted lung development and an abundance of pro-inflammatory cells and proteins in the lung^40,41^. There is evidence that the lung inflammation in BPD is reflective of a systemic pro-inflammatory state^42,43^, yet the specific immune perturbations associated with disease remain poorly understood. The incidence of BPD is inversely proportional to gestational age at birth, and most extremely preterm infants develop BPD^44^. However, even controlling for gestational age, preterm infants can develop widely different severities of BPD. Infants who develop severe BPD (Grade 3) have very different clinical courses than those with moderate BPD (Grade 2) and the former requires prolonged invasive mechanical ventilation with disproportionate impact on long-term health^45^. Thus, to explore how severe BPD is associated with changes in immune development, we focused on a gestational-age controlled subset of preterm infants born less than 28 weeks GA and defined severe or moderate BPD by the level of respiratory support required by each infant at 36 weeks cGA^45^ (**Figure 3A**). The overall immune landscape of preterm infants who developed severe BPD was progressively more different each month after birth from those with moderate BPD in PCA space (**Figure 3B**). This increasing immune divergence was driven by differences in neutrophils, CD4^+^ T cell sub-populations and some changes in NK cells and plasmacytoid DCs (**Supplemental Figure 3A**). To further interrogate how immune cells changed over time between these two BPD groups, we constructed a mixed linear model for each cell population. Neutrophil frequencies remained steadily elevated in severe BPD while they progressively decreased in moderate BPD, contributing to the overall divergence seen each month (**Figure 3C**). Dexamethasone is frequently used in infants with BPD and can increase the circulating neutrophil pool via neutrophil demargination. Dexamethasone is typically given to infants in short courses to minimize side effects, and nine samples in our study were collected during active dexamethasone exposure. The samples collected while these infants were receiving dexamethasone had no significant difference in neutrophil frequencies compared to samples from infants who were not receiving the therapy (**Supplemental Figure 3B**), suggesting that the observed divergence in neutrophil proportion was not due to active dexamethasone administration.

**Figure 3:**
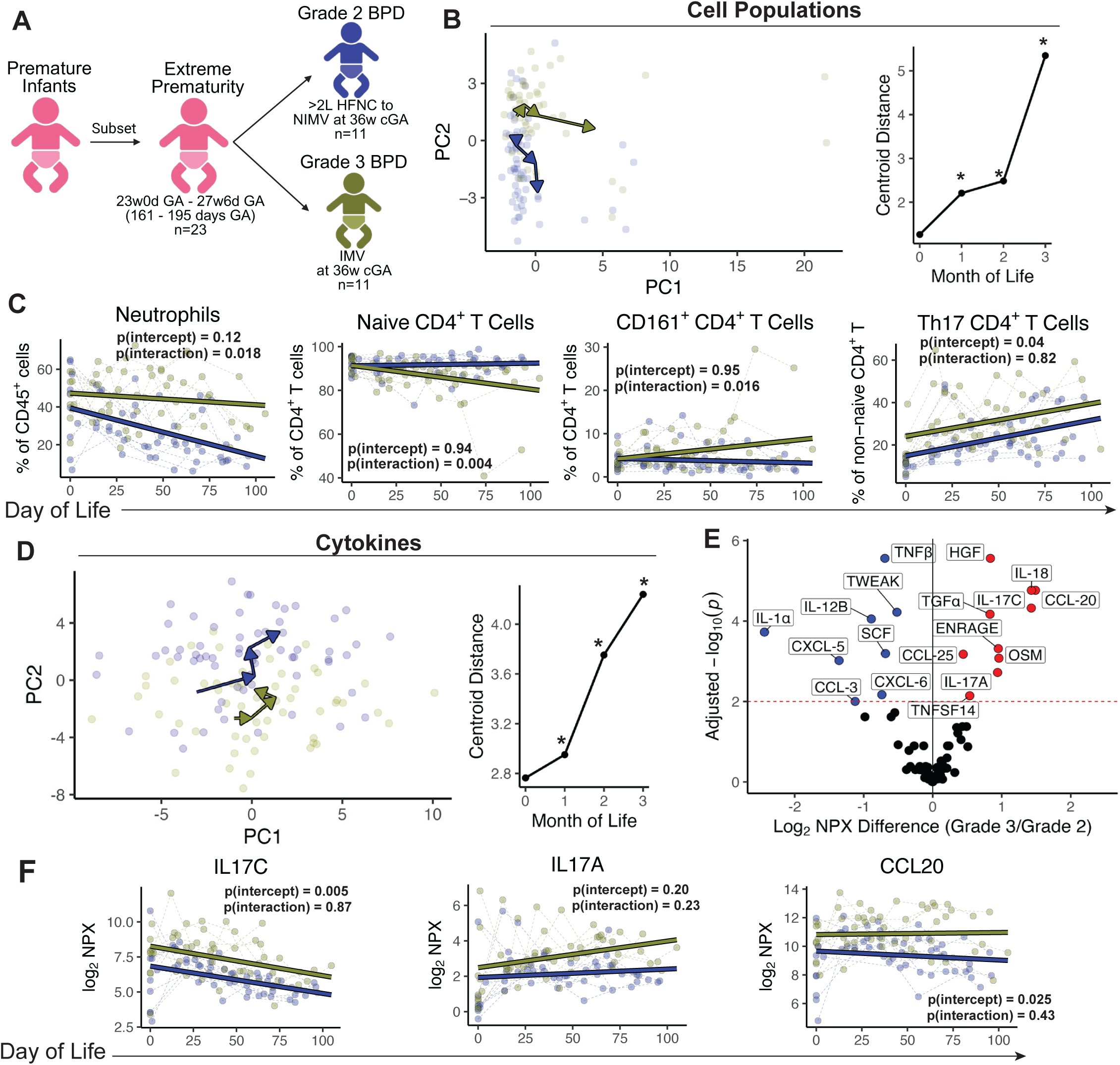
Severe preterm lung disease is associated with a divergent Th17 T cell signature. A) Schematic depicting the infants analyzed for this figure. One infant in the extreme prematurity subset developed Grade 1 BPD and was not included in further analyses in this figure. HFNC = High flow nasal cannula, NIMV = non-invasive mechanical ventilation, IMV = invasive mechanical ventilation. B) PCA of all cell populations in postnatal samples from infants with Grade 2 BPD and Grade 3 BPD. Arrows point to a centroid at each sequential month of life. C) Euclidean distances between the two centroids at each month of life. *=*p*<.01 via PERMANOVA. C) Mixed linear regressions of neutrophils, naïve CD4^+^ T cells, CD161^+^ CD4^+^ T cells and Th17 CD4^+^ T cells. Intercept p-value (representing categorical difference at birth) and interaction p-value (representing divergence over time) shown for each regression. D) PCA of all cytokines in postnatal samples from infants with Grade 2 BPD (blue) and Grade 3 BPD (green). Euclidean distances between the centroids at each month of life. *=*p*<.01 via PERMANOVA. E) Volcano plot demonstrating cytokines that differ between groups. Cytokines increased in Grade 3 BPD are on the right. F) Mixed linear regressions of IL-17C, IL-17A and CCL20.

Infants with severe BPD also had a progressive decline in the frequency of naïve CD4^+^ T cells, suggesting greater CD4^+^ T cell activation over time (**Figure 3C**). There was also a progressive increase in the frequency of CD161^+^ CD4^+^ T cells (**Figure 3C**). CD161^+^ CD4^+^ T cells are notable as they are primed to secrete Th17-related cytokines and give rise to non-naïve Th17 T cells^35,36^. In examining the polarization phenotype of all non-naïve CD4^+^ T cells, the non-naïve CD4^+^ T cells of infants with severe BPD were biased towards a Th17 (CCR4^+^, CCR6^+^, CXCR3^-^) phenotype, but not a Th1 (CCR4^-^, CCR6^-^, CXCR3^+^) or Th2 (CCR4^+^, CCR6^-^, CXCR3^-^) phenotype (**Figure 3C**, **Supplemental Figure 3C**, and **Supplemental Figure 3D**). It was notable that whereas neutrophil, naïve CD4^+^ T cell, and CD161^+^ CD4^+^ T cell frequencies were similar early and diverged over time, Th17 cells were elevated in severe BPD infants from birth and this difference persisted throughout the time in the NICU.

One of the primary functions of Th17 cells is to promote neutrophil responses at sites of inflammation such as the lung through production of Th17-related cytokines and chemokines^46,47^. The overall pattern of plasma cytokines and inflammatory mediators diverged between infants with severe BPD and infants with moderate BPD (**Figure 3D**). Among the cytokines increased in severe BPD were IL-17A, IL-17C, and the ligand of the characteristic Th17 chemokine receptor CCR6, CCL20 (**Figure 3E**). Other pro-inflammatory Th1-related cytokines, such as IL-12β, lymphotoxin, and IL-1α were lower in infants with severe BPD, highlighting a bias to a Th17 and not a Th1 inflammatory state. Mixed linear model analysis demonstrated that infants with severe BPD had elevated IL-17C and CCL20 at birth and throughout their time in the NICU (**Figure 3F**). IL-17A did not reach statistical significance in the linear model but trended in the same direction. Together, these data show that the increased neutrophil frequency observed in preterm infants with severe BPD was associated with early-life alterations in Th17-biased CD4^+^ T cells and Th17-related cytokines, and that these differences may already be present at birth.

### Diverse early-life infections are associated with robust CD8^+^ T cell responses

Although the severity of BPD was associated with some of the postnatal immune perturbations in this cohort, severe BPD alone could not account for all immune changes. Notably, the frequency of naïve T cells was similar in infants with severe versus moderate BPD, yet four infants had a sudden and robust decrease in the frequency of naïve CD8^+^ T cells during their time in the NICU (**Figure 2C**). For three of the infants, this change in naïve CD8^+^ T cell abundance occurred within days of a new onset laboratory-confirmed systemic infection, including blood culture-confirmed *Staphylococcus epidermidis*, blood culture-confirmed methicillin-resistant *Staphylococcus aureus* infection, and blood PCR-confirmed cytomegalovirus (CMV) infection. In one infant, P36, this change in CD8^+^ T cells occurred more than a month after both a diagnosis of necrotizing enterocolitis (NEC) infection and blood culture-confirmed *Staphylococcus hominis* infection. In each of these four infants, the typical distribution of 85-99% CD45RA^+^CD27^+^CCR7^hi^ naïve-phenotype CD8^+^ T cells changed abruptly to ∼50-80% of the circulating CD8^+^ T cells displaying an activated or antigen-experienced differentiation state based on CD45RA, CD27, and CCR7 (**Figure 4A**). This non-naïve CD8^+^ T cell population was comprised mostly of EM-type cells (CD45^−^ CD27^+^ or ^−^). Although individual distributions of CD8^+^ T cell subsets varied, there were some general patterns over time, with effector memory 1 cells (EM1, CD45RA^-^ CD27^+^ CCR7^-^) expanding first, followed by effector memory 3 cells (EM3, CD45RA^-^ CD27^-^, CCR7^-^), and then, often weeks later, the emergence of terminally differentiated T effector memory re-expressing CD45RA cells (TEMRA, CD45RA^+^, CD27^-^, CCR7^-^) and CD57^+^ CD8^+^ T cells (**Figure 4A**). In each infant, the changes in CD8^+^ T cell subset distribution persisted at least until each infant exited the study, over a month after the associated infection had resolved.

**Figure 4:**
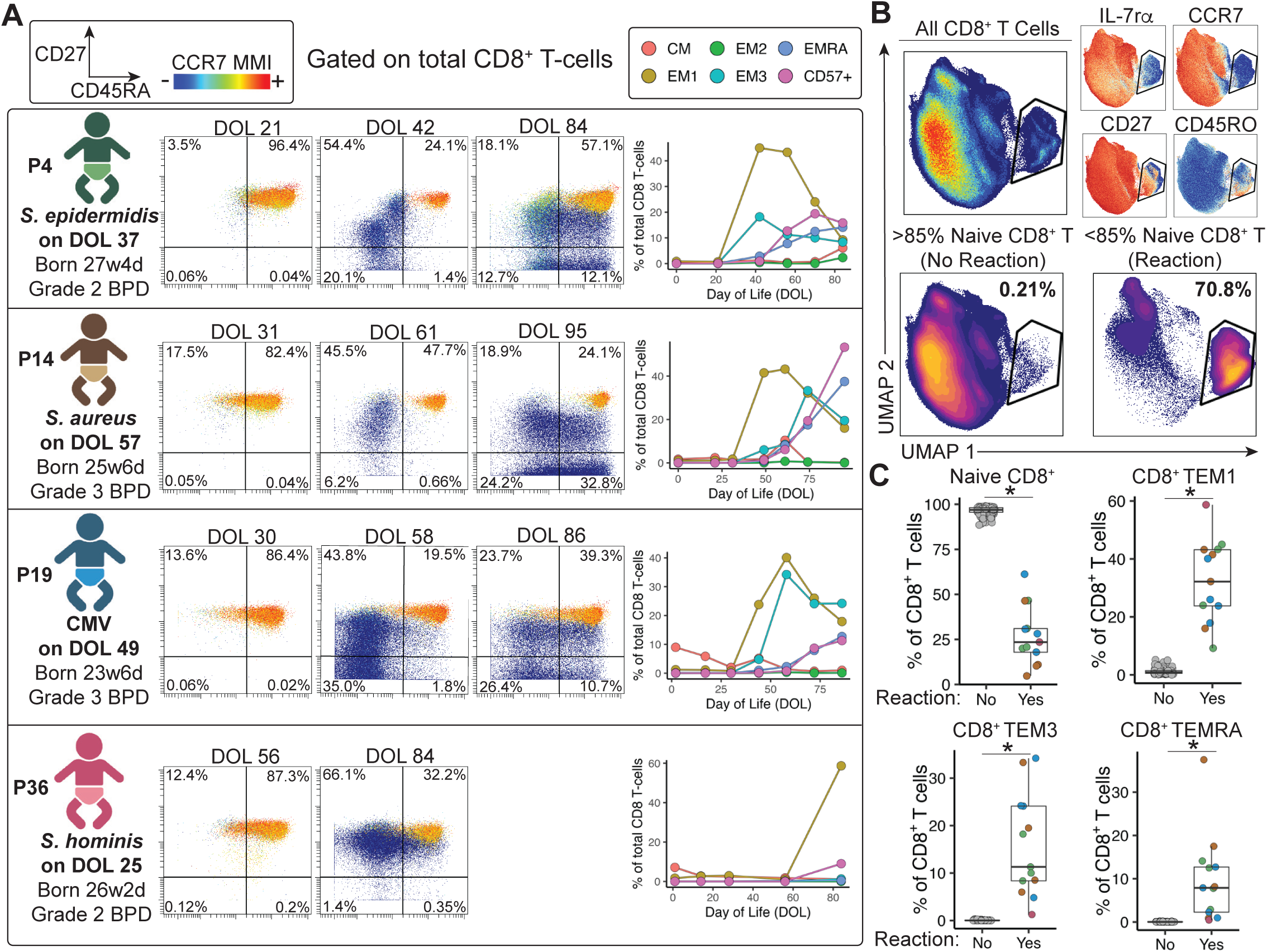
Bloodstream infection is associated with a sudden, massive, and persistent CD8^+^ T cell reaction. A) Schematic representing the progression of the total CD8^+^ T cell population of four infants in the study. Sequential CyTOF plots gated on all CD8^+^ T cells shown for each infant. The day of life each sample was collected is denoted above each plot. For each CyTOF plot, X-axis = CD45RA, Y-axis = CD27, Z-axis (color) = CCR7. Overview plot to the right of each sequence represents specific CD8^+^ T cell sub-populations as they change over time including CM, EM1, EM2, EM3, EMRA, and CD57^+^ cells as a percent of the total CD8^+^ T cell population. B) UMAP of all CD8^+^ T cells derived from all samples. Expression of specific markers, IL-7Rα, CCR7, CD27, and CD45RO overlayed on the CD8^+^ T cell UMAP for reference. Density UMAP of all samples with >85% naïve CD8^+^ T cells (denoted ‘No Reaction’) and <85% naïve CD8^+^ T cells (denoted ‘Reaction’). C) CD8^+^ T cell sub-populations found in samples with and without a reaction. *<p=0.05 via BH-adjusted Wilcoxon test.

To investigate these neonatal CD8^+^ T cell responses in more detail, we defined any samples from infants in whom the fraction of CD8^+^ T cells fell below 85% naïve as having had a ‘reaction’. Once an infant first developed a reaction, all subsequent samples also fell below 85% naïve CD8^+^ T cells. In samples from these infants, over 70% of the CD8^+^ T cell population shifted to CCR7^lo^, CD27^lo^, CD45RO^hi^ and IL7Rα^lo^ (**Figure 4B**). In some subjects at some time points, less than 10% of the CD8^+^ T cell population remained naïve. CD8^+^ T cell EM3 and TEMRA populations that became the predominant populations in these infants were nearly undetectable in infants that did not have a reaction (**Figure 4C**).

The microbes associated with these reactions were, in some cases, unexpected. While one infant (P19) experienced an early-life infection with CMV, perhaps consistent with the induction of a CD8^+^ T cell response, two other infants (P4 and P14) experienced bacterial infections (*S. epidermidis* and *S. aureus*), two infections not typically associated with robust CD8^+^ T cell responses. A fourth infant (P36) had no proximal infection detected at the time of the CD8^+^ T cell response but had experienced a *S. hominis* infection at an earlier time point. Although it is possible that infections with clinically undetected intracellular pathogens also occurred concurrently with the confirmed bacterial infections, these events suggest a role for CD8^+^ T cells during both early-life intracellular and extracellular infections in preterm infants. There were no infants without a bloodstream infection who developed a CD8^+^ T cell reaction and four of seven infants in this study with a confirmed bloodstream infection developed a CD8^+^ T cell reaction. In sum, these observations suggest that robust and dynamic T cell changes may indicate infections even before, or in the absence of, pathogen detection by routine clinical infectious disease testing.

### The CD8^+^ T cell reaction to early-life infection is associated with CD4^+^ T cell, ψ8 T cell, and plasma inflammatory cytokine changes

We next investigated whether there were other immune changes in infants with an infection-associated CD8^+^ T cell reaction. Indeed, there was induction of CD45RO^hi^, CCR4^hi^, CD45RA^lo^, CCR7^lo^ CD4^+^ T cell responses and an increase in EM3 and TEMRA CD4^+^ T cell populations in infants with a CD8^+^ T cell reaction (**Figure 5A** and **Figure 5B**). CD8^+^ T cell reaction samples also had a reduced frequency of naïve CD4^+^ T cells, and increased frequency of EM3, and TEMRA subsets (**Figure 5B**). Although EM1 cells made up the largest population of non-naïve CD4^+^ T cells, these cells were not increased in reaction samples, likely reflecting ongoing EM1 cell elevation over time in non-reaction samples such as those from infants with severe BPD. However, reaction samples had a sharp increase in CCR6^−^CCR4^−^CXCR3^+^ Th1 cells and CCR6^+^CCR4^+^CXCR3^−^ Th17 cells, and a strong decrease in CCR4^+^CCR6^−^CXCR3^−^ Th2 cells (**Figure 5C** and **Figure 5D**). In addition to this CD4^+^ T cell response, there was also a ψ8T cell response in the infants with infection-associated CD8^+^ T cell reactions with a shift to CD27^lo^, CCR7^lo^, CD161^lo^, and IL7rα^lo^ ψ8T cells (**Figure 5E**). Using analogous marker subsets for αβT cells to look at ψ8T cell phenotype, there was little change in ψ8 T EM1-like cells, but the frequency of EM3-like and TEMRA-like ψ8T cell populations were increased (**Figure 5F**). Thus, whereas ψ8T cell subpopulations change in most infants after birth with a gradual increase in CD161^+^ and non-naïve ψ8T cells, only infants with an infection-associated CD8^+^ T cell reaction have ψ8T cell responses resulting in differentiation into subpopulations analogous to αβT EM3 and TEMRA populations.

**Figure 5:**
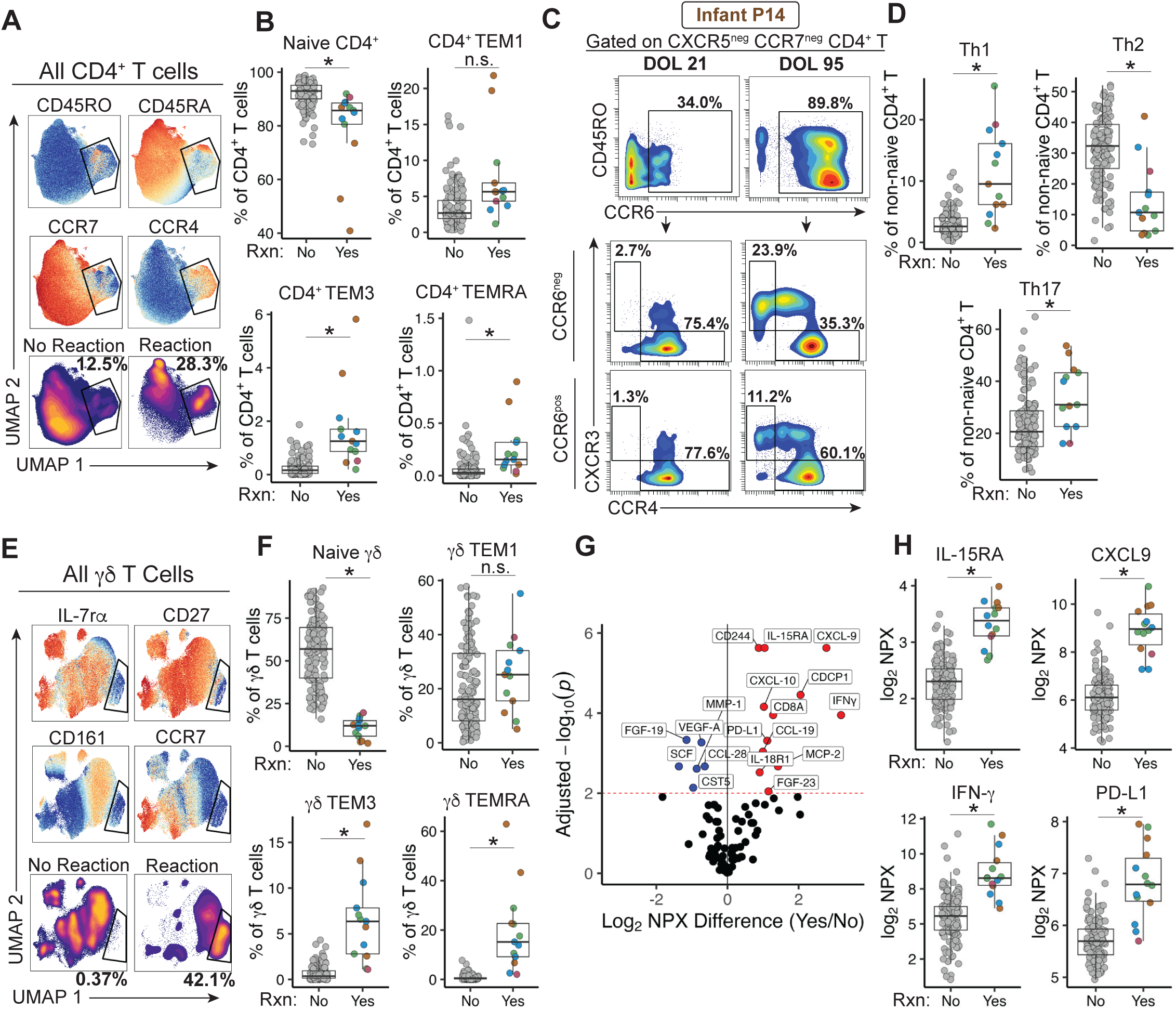
CD4+ T cells, γδ T cells, and plasma cytokines shift along with the infection-associated CD8+ T cell reaction. A) UMAPs of CD4^+^ T cells from all samples showing the expression of different surface markers, and then UMAPs representing the density distribution of samples identified as having a CD8^+^ T cell reaction or not. B) Different CD4^+^ T cell sub-populations (EM1, EM2, EM3, and EMRA) found in samples with and without a T cell reaction. C) Representative CyTOF plots showing the distribution of Th subpopulations of non-naïve CXCR5^-^, CCR7^-^T cells from one sample before a reaction and one sample after a reaction from infant P14. D) Average Th1, Th2, and Th17 phenotypes found among samples with and without a reaction. E) UMAP of all γ8T cells derived from samples identified as having a CD8^+^ T cell reaction or not. F) Different γ8T cell sub-populations found in samples with and without a reaction. G) Volcano plot showing the difference in plasma cytokines from all samples with and without a reaction. Cytokines increased in reaction samples on right. H) Specific plasma cytokines IL15-RA, CXCL9, interferon-γ, and PD-L1 shown. *<p=0.05 via BH-adjusted Wilcoxon test.

These T cell responses in infants with infections were also associated with many changes in plasma cytokines and inflammatory proteins, including an increase in many proteins related to CD8^+^ T cell and CD4^+^ Th1 responses, including IFNγ, IL15Rα, CXCL9, CXCL10 (a ligand, along with CXCL9, for the Th1 receptor CXCR3), soluble IL-18R1, and soluble PD-L1, together with lower FGF-19, VEGF-A and others (**Figure 5G** and **Figure 5H**). Thus, in a subset of preterm infants in this study, systemic viral or bacterial infection was associated with robust CD8^+^, CD4^+^, and ψ8 T cell responses and associated circulating inflammatory mediators leading to durable changes in the circulating adaptive immune compartment for at least several weeks after resolution of infection.

### The infection-associated T cell responses are oligoclonal and progressive

Given such a robust shift in T cell differentiation state in the blood of certain infants with infection, we next investigated whether this event represented a non-specific polyclonal T cell response, as has been reported in neonatal T cells^48,49^, or was more oligoclonal suggesting an antigen-driven response. We isolated DNA for T cell receptor (TCR) sequencing from time points before and after an infection and T cell response from three of the infants with a reaction. The blood samples from infant P4 were not available for this analysis. We also performed TCR sequencing for two other age- and BPD-matched infants who did not develop a bloodstream infection. TCR clonality increased progressively in each infant after infection (**Figure 6A**) and the TCR pool became clonally dominated soon after the infection with increasing clonality over the following month (**Figure 6B** and **Figure 6C**). Prior to infection, or in infants with no infection, there was limited to no TCR expansion. In infants with infection, although some expanded clones were not captured at every time point, there were expanded clones that were shared between serial time points (**Figure 6B**). Thus, the T cell responses observed in preterm infants with infection were associated with robust TCR oligoclonal expansion.

**Figure 6:**
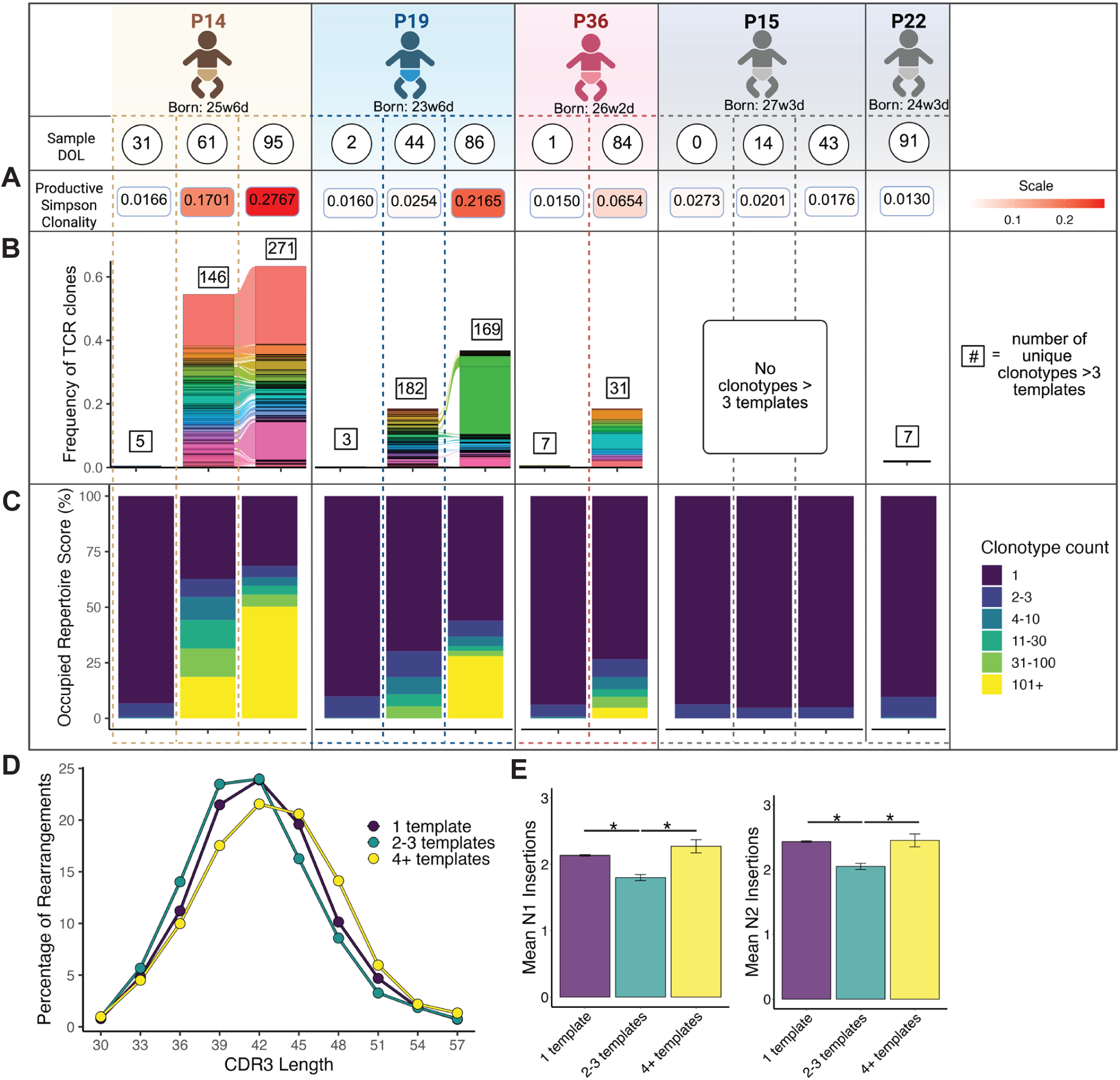
Massive first-in-life T cell reaction is oligoclonal. TCR-seq analysis of select samples from infants with a T cell reaction. Information from each sample can be seen as a sub-column under the infant from which it was derived. A) The productive Simpson clonality index of each sample. B) The frequency of specific TCRs that were found to have greater than three clonotypes within the sample. Alluvia connecting the colors between samples represent the same TCR sequence that was found in both samples. C) The occupied repertoire score, which is the percent of the sample that is dominated by TCRs of a given clonotype count. D) The CDR3 length of TCRs within each clonotype count bin. E) The mean number of N1 and N2 insertions among TCRs within each clonotype count bin. * = p<0.05 via ANOVA.

We next asked whether the highly expanded TCR clones varied in their complementarity-determining region 3 (CDR3) length or number of N insertions compared to unexpanded or mildly expanded clones. In general, preterm CDR3 lengths are shorter, with fewer N-insertions, compared to mature adult TCRs^10^. In infants with infection-associated expanded TCR clones, the CDR3 lengths of expanded clones tended to be longer than those of less expanded TCR clones (**Figure 6D**). The number of N-insertions in TCR clones with four or greater copies was also increased compared to clones with two or three copies, though TCRs represented by only a single TCR copy had similar N-insertions to the highly expanded clones (**Figure 6E**). TCR clones with two or three copies can be found with similar frequency in all the samples sent for TCR analysis, whereas TCR clones with greater than four copies were uniquely found in samples with infection reaction. Thus, despite a restricted early-life TCR repertoire^10,50^, preterm infant infection with CMV or extracellular bacteria in the NICU was associated with robust TCR clonal expansion and persistent reshaping of the TCR repertoire.

## Discussion

Recent work has begun to dissect the unique dynamics of the human infant immune system^16,51,52^. A major gap in our understanding of early-life human immune development, however, is in the setting of preterm birth, where immune development is perturbed by postnatal stressors and clinical disease. To address this question, we developed a high-dimensional longitudinal immune profiling approach to interrogate the trajectory of the preterm immune system during postnatal life in the NICU. Using this approach, we captured early-life immune development changes in preterm infants highlighting rapid changes in neutrophils, T cells, and inflammatory mediators when infants were forced to adapt to birth prior to full gestation. In addition, we captured the impact of two common clinical comorbidities of prematurity on immune development. First, infants who developed severe BPD were characterized by a progressively Th17-dominated immunologic signature. Second, some infants who developed viral or extracellular bacterial infection had a robust oligoclonal T cell response dominated by CD8^+^ T cells, along with CD4^+^ T cell responses, ψ8T cell responses, and type 1 cytokine changes. Many of these durable immune changes were continuing to evolve when the observational period of the study ended and thus may have immune imprinting effects that extend beyond this period of clinical observation.

Th17 CD4^+^ cells have a central role in many autoimmune and inflammatory disorders^53^, and may also contribute to pathophysiologic changes in preterm lungs^54^. Neonatal CD4^+^ T cells readily undergo Th17 polarization in a manner that is inversely related to gestational age^55^. Our observations that the proportion of CD161^+^ CD4^+^ T cells inversely correlates with gestational age in infant blood, a finding also seen in studies of cord blood^31,36^, are consistent with findings of a Th17 bias in the early-life immune system. It is worth noting that the risk of severe BPD also inversely correlates with gestational age, and thus extremely preterm infants predisposed to Th17 polarization also have a developmental lung state most at risk of severe BPD. The data here further show that infants with severe BPD have progressive increases in Th17 CD4^+^ T cells, circulating Th17 cytokines, and neutrophils over time, all characteristic of a Th17 signature. Thus, we speculate that extremely preterm infants who activate CD4^+^ T cells early in life have a bias towards inducing and then reinforcing the Th17 program, thereby increasing the likelihood of neutrophilic inflammation which could damage an already immature lung architecture. Indeed, several studies support this notion. For example, in mouse models of neonatal BPD-like lung inflammation, blockade of the Th17 system reduces the degree of inflammation and damage in the lung^56,57^. Additionally, histologic chorioamnionitis at birth, a risk factor for developing BPD, is associated with an increase in Th17 cells in the cord blood, implicating type 17 responses in preterm inflammation^58,59^. Current methods of reducing inflammation in BPD, such as dexamethasone, have numerous side effects that make them difficult to use as an ongoing therapy in infants^60^. The IL-17 axis offers a more specific potential target for intervention in BPD, and there exist multiple, safe, approved therapies that target the Th17 system^61^. Whether blockade of the Th17 axis is a viable anti-inflammatory approach to reduce the risk or severity of BPD warrants further study.

In addition to severe BPD, systemic infection in a subset of infants in our cohort was associated with unique and profound changes to the T cell pool. Indeed, in part because our scavenged blood design allowed us to frequently profile preterm infants, we captured multiple robust first-in-life T cell reactions in real-time. These findings add to the evidence that the preterm immune system, despite its restricted T cell repertoire and resistance to certain inflammatory stimuli, can be highly reactive and proinflammatory, mounting vigorous adaptive immune responses in some settings of infection^4,5^. Robust CD8^+^ T cell activation in infants has been observed following infection with several viruses, including acquired CMV^62^, congenital CMV^63^, and HIV^64^. These studies reported that durable alterations in the CD8^+^ T cell compartment could be found even 12 months after infection in term infants^62^. These new data in preterm infants, in addition to identifying coordinated CD8^+^ T cell, CD4^+^ T cell, ψ8 T cell, and plasma protein changes, suggest extensive oligoclonal T cell activation might also be a feature of severe bacterial infection, in addition to viral infection, in neonates. Indeed, studies in rodents have demonstrated that neonatal CD8^+^ T cells are critical for controlling extracellular bacterial infections^65^. Moreover, neonatal T cells have the ability to secrete CXCL8, a neutrophil-attracting chemokine, whereas adult T cells do not^13^. These observations suggest that neonatal T cells may play a larger role in conditions where neutrophil activity is needed, such as bacterial infections. There were no concomitant viral infections diagnosed in the infants with a laboratory-confirmed bacterial blood culture. It remains possible, however, that an undiagnosed concurrent viral infection was present in these settings. It is also notable that many of the infants in our study who reached two months of age likely received the full complement of vaccinations around this time. Although we could not coordinate sample acquisition around the timing of these vaccinations, immunologic reactions to vaccination were not obvious in our dataset, suggesting that infection, rather than early-life vaccination, was needed to trigger these robust adaptive immune responses. Regardless of the trigger, the physiologic relevance of such reactions, including how children who experienced these early-life infections and immune responses might respond to future infections or non-infectious immune challenges, warrants further study. Moreover, the differentiation pattern and T cell subset distributions in these preterm infants are somewhat comparable to those found in the healthy adult CD8^+^ T cell compartment^66^, suggesting that this sudden shift to an expanded, differentiated, and TCR matured CD8^+^ T cell pool may be an event that occurs in all humans at some point during life. If true, how and when this event occurs may have significant immunologic consequences.

There are likely many other immunologic or clinical conditions, not captured in this cohort, that are associated with durable changes in the immune trajectory. Detailed information for each preterm infant, such as whether the infant was born due to maternal pre-eclampsia or preterm labor, or whether they were born in the setting of histologic chorioamnionitis, could not be captured in the current cohort due to limitations on collected clinical data. Even after birth, there are other major comorbidities of prematurity, including NEC, severe interventricular hemorrhage, or severe retinopathy of prematurity which were not captured with enough power to make biologic conclusions here but warrant future investigation given that they may be associated with their own unique immune perturbations^67,68^. Preterm infants are also exposed to numerous stressors in the NICU that we did not track in this study. One exposure that is of particular interest is commensal microbes. *Olin et al.* reported that infants with microbial dysbiosis have greater interindividual diversity compared to infants with a healthy microbiome, and suggest that otherwise stereotypic immune development may be altered in these infants^16^. Dysbiosis has been associated with a number of immunologic changes early in life that can precede diagnosis of disease later^17,25,69^. In animal models, rodents with dysbiosis have increased Th17 cell-driven asthma that persists even after normalizing their microbiome^70^. When considered with our findings of a progressive Th17 bias in BPD, a condition in which asthma-like airway hyperreactivity is widely prevalent^71^, this set of work suggests that the infant microbiome may play a key role in predisposing infants to disease-specific immunologic skewing. While the current work provides a foundation for interrogating the preterm infant immune system and dynamic trajectory, there are numerous unmeasured factors that may drive immunologic diversity that warrant further study.

Overall, these data document major immunological changes associated with common clinical events of prematurity and suggest lasting consequences for immune development. Moreover, we reveal that different early-life comorbidities can change immune trajectories in unique ways. Thus, while each preterm infant immune system will ultimately become “adult”, the path it takes to get there can be shaped and potentially re-wired by factors such as infection or severe lung disease. Each of these unique trajectories may reflect ongoing illness, as with evolving severe BPD, or past illness, as with systemic infection. Defining how these early-life immunologic trajectories ultimately influence the development of childhood and even adult immune health will be an important future goal.

## Methods

### Study Overview

Preterm infants admitted to the Hospital of the University of Pennsylvania Intensive Care Nursery were considered eligible for the study if they met criteria of born between 23 weeks and 0 days and 32 weeks and 6 days. Infants were excluded from the study if they had any known genetic diagnosis at birth, any severe congenital anomalies, or were born in the setting of human immunodeficiency virus (HIV). Sex was not considered when deciding enrollment, and we ultimately enrolled a similar number of male and female infants in the study. After enrollment, residual unused blood from the first CBC of life, often sent by clinicians in the first 24 hours of life, was obtained from the clinical lab and deidentified. Whole blood was collected from the HUP clinical lab 24-48 hours after it had completed CBC analysis and was to otherwise be discarded. A minimum of 12 days was required between serial sample collection for each infant. Blood was recurrently obtained based on the availability of samples until an infant was discharged or met 40 weeks cGA. All blood was collected in K2-EDTA vacutainers. Samples were stored at room temperature in the clinical lab until processing.

### Clinical Data Collection

Selected clinical data as approved by the Institutional Review Board (IRB) were obtained for each infant including age, sex, and weight at birth, time and results of each CBCs sent, the diagnosis of major comorbidities as would appear in the electronic medical record, time and results of blood, urine and cerebrospinal fluid infectious testing sent on each infant, and the start and end dates of dexamethasone or hydrocortisone exposure. All clinical data was stored on a secured REDCap database contained within the University of Pennsylvania system.

### Plasma cytokine analysis

Fifty μL of whole blood was centrifuged at 2000g for 15 minutes, and plasma removed and stored at -80°C until analysis via the Olink target 96-inflammation kit. Data were reported as normalized protein expression values (NPX). The NPX values for two batch runs were normalized using bridging samples present in both analyses. Cytokines were excluded from subsequent analysis if >25% of the samples analyzed were below the limit of detection for the assay or the cytokine failed quality control metrics. Eight cytokines were ultimately excluded for these technical reasons, resulting in 88 included cytokines.

### CyTOF analysis

Up to 300μL of whole blood was stained with the MaxPar Direct Immunophenotyping Assay (MDIPA) lyophilized antibody cocktail (Standard BioTools Inc.) for 30 minutes at room temperature^72^. The assay requires 300μL of whole blood. If the available blood volume was less than 300μL, heat-inactivated pooled human AB serum was added to meet 300μL. After 30 minutes, 420μL of PROT1 proteomic stabilizer buffer (SmartTube Inc, San Carlos, CA) was added, followed by 10 minutes incubation at room temperature. Samples were then stored at -80°C until batched analysis. Samples were thawed and RBC lysis was performed using Thawlyse buffer (Smart Tube Inc) as per manufacturer’s recommendation. 125nM iridium (Cell-ID™ Intercalator-Ir, Standard BioTools Inc) in MaxPar Fix Perm buffer (Standard BioTools Inc) was then added and samples were stored at 4°C until further processing for acquisition. A batch control sample, prepared by staining a single large batch blood sample drawn from a healthy adult donor, was included in each batch of sample acquisition. Samples were washed twice with MaxPar Cell Staining Buffer (Standard BioTools Inc), followed by two washes with Cell Acquisition Solution+ (CAS+, Standard BioTools Inc), filtered through 35μm nylon mesh and loaded on CyTOF XT mass cytometer (Standard Biotool Inc) for data collection. Daily instrument QC and tuning was performed using EQ^TM^ Six (EQ6) element calibration beads (Standard BioTools Inc). EQ^TM^ Four (EQ4) element calibration beads (Standard BioTools Inc) were added at a 10% v/v to the samples that were suspended at a concentration of 1 million per mL in CAS+ buffer. The CyTOF® Software v9.1, data acquisition software, used signals from EQ4 bead channels to normalize data from analyte channels. Cells were injected into the detector at ∼500 events per second and at a flow rate of 30μL per minute. Manual gating of immune cell populations was performed using OMIQ software (Dotmatics) to identify immune populations for analysis. All raw CyTOF data were arcsin scaled prior to gating. All full gating scheme is provided in **Supplemental Figure 1**.

### TCR sequencing

DNA was isolated from fixed immune cells remaining after CyTOF analysis with the PureLink Genomic DNA Mini Kit (Invitrogen). Briefly, cells were de-crosslinked and incubated with proteinase K overnight. The next day samples were incubated with RNAase and spun into a DNA collection tube, washed, and eluted. DNA purity and quality was assessed via NanoDrop (Thermo Fisher). Isolated DNA from each sample was sent for TCR sequencing (Adaptive Biotechnologies). Initial data were analyzed using the ImmunoSEQ Analyzer 2.0 toolkit and exported for further analysis and plotting in R.

### Statistics

All deidentified data were exported from REDCap and stored in secure Microsoft Excel data frames and analyzed via R. Correlations for immune features over gestation were performed using a Spearman’s rank coefficient. All PCA plots generated were scaled. PCA plots were statistically analyzed via PERMANOVA tests using the *vegan* package in R. All linear mixed models were generated and analyzed using the *lme4* package in R. To construct a fixed-effects regression line as used in **Figure 2**, a prediction grid was constructed spanning the range of observed cGA days and predicted the fixed effect estimates from the fitted mixed model with the random (infant-specific) effect set to zero. TCR data were plotted using the *ggalluvial* package. All *p* values were BH corrected, and thresholds of significance are reported in each figure. All other data was plotted and statistically analyzed using R packages, including *ggplot2, dplyr, tidyr, ggpubr, reshape2,* and *stats*. Summary figures were created using BioRender.

## Ethics and Study Approval

This study was reviewed and approved by the University of Pennsylvania Institutional Review Board (IRB). As this study used residual blood from samples collected for clinical purposes and had no patient-facing interactions, this study was approved with a waiver of consent, which limited potential selection bias. Each category of clinical information gathered from the medical record was outlined and reviewed by the IRB prior to collection and all data was stored in a secure de-identified REDCap database.

## Author Contributions

B.A.F. and E.J.W. conceived of and designed the research study. B.A.F., E.J.W. and K.L. wrote and sponsored the IRB protocol. B.A.F., M.M., S., A.P., Z.M., C.D., acquired the data from collected samples. B.A.F., D.M., M.M., A.R.G., E.J.W., developed the data analysis approaches. B.A.F. and E.J.W. wrote the manuscript. All authors critically reviewed the manuscript.

## Data Availability

Data in the present study are contained in the main manuscript or supplemental information or are available upon reasonable request to the authors.

## Acknowledgements

We would like to thank the Hospital of the University of Pennsylvania clinical laboratory for assisting with efforts in obtaining these residual samples, members of the University of Pennsylvania Immune Health team for assisting with sample transport, and the members of the Wherry lab and Immune Health for their critical discussions of the data and manuscript. BAF was supported by the Thrasher Early Career Research Award, the American Association of Pediatrics (AAP) Marshall Klaus Award, and the Division of Neonatology and the Children’s Hospital of Philadelphia. This work was supported by NIH grants AI155577, AI115712, AI117950, AI108545, AI082630, AI149680, HL145754 (to E.J.W.), Parker Institute for Cancer Immunotherapy (to E.J.W.), and The Mark Foundation (to E.J.W.), the Arguild Foundation and philanthropic support to the UPenn Immune Health Innovation fund.

