## Supplemental Figures for "Perturbation of the Preterm Human Immune System in Early Life"

**Supplemental Figure 1: Gating Strategy**. Cells in bold were analyzed as a proportion of total CD45+ cells.

Cells underlined were analyzed as a proportion of the parent cell in bold above it.

> CD45+ Cells (CD45+, DNA+)
>> Granulocytes (CD66b+, CD45 lo-mid)
>>> **Neutrophils** (CD294-, CD16hi)
>>> **Eosinophils** (CD294+, CD16lo)
>> Mononuclear Cells (non-Granulocytes)
>>> Non-monocyte, non-NK cell (CD14-, CD56-)
>>>> Non-T (CD3-, IL3R-)
>>>>> **B cells** (CD19+, CD20+)
>>>>>> Naïve B cell (IgD+, CD27-)
>>>>>> IgD negative Memory B cell (CD27+, IgD-)
>>>>>> IgD positive Memory B cell (CD27+, IgD+)
>>>>> CD20 negative (CD20-)
>>>>>> **Plasmablast (CD38+, CD27+)**
>>> Non-T cell, non-B cell (CD3-, CD19-)
>>>> Non-NK cell (CD56-, CD20-)
>>>>> **Monocytes (CD11c+)**
>>>>>> Classical Monocytes (CD14+, CD16-)
>>>>>> Intermediate Monocytes (CD14+, CD16+)
>>>>>> Non-Classical Monocytes (CD14-, CD16+)
>>>>> **Basophils (IL-3R+, HLA-DR-)**
>>>> Non-Monocyte (CD14-, CD20-)
>>>>> Dendritic Cells (HLA-DR+, CD56-)
>>>>>> **Plasmacytoid Dendritic Cells (CD11c-, IL-3R+)**
>>>>>> **Classic Dendritic Cells (CD11c+, IL3R-)**
>>>>> **NK Cells (CD56+, HLA-DR-)**
>>>>>> CD56 bright NK Cells (CD56 hi, CD16-)
>>>>>> CD56 dim NK cells (CD56 lo-mid)
>>>>>>> Early NK Cells (CD57-)
>>>>>>> Late NK Cells (CD57+)
>>> T cells (CD3+, CD19-)
>>>> **NK T cells (CD56+)**
>>>> **γδ T cells (TCR γδ+)**
>>>> ab T -cells (TCR γδ-)
>>>>> **CD8 T cells (CD8+, CD4-)**
>>>>> **CD4 T cells (CD8-, CD4+)**
>>>>> **Double positive T cells (CD8+, CD4+)**
>>>>> **Double negative T cells (CD8-, CD4-)**
--The following applies to CD8+ T cells, CD4+ T cells, and γδ T cells each--
>>>>>> CD161+ T cells (CD161+)
>>>>>> CD57+ T cells (CD57+)
>>>>>> CD27+ T cells (CD27+)
>>>>>>> Effector Memory 1 T cells (CCR7-, CD45RA-)
>>>>>>> CCR7+ T cells (CCR7+)
>>>>>>>> Central Memory T cells (CD45RO+, CD45RA-)
>>>>>>>> Naïve T cells (CD45RA+)
>>>>>> CD27- CD45RA- T cells (CD27-, CD45RA-)
>>>>>>> Effector Memory 2 T cells (CCR7+)
>>>>>>> Effector Memory 3 T cells (CCR7-)
>>>>>> T-effector memory re-expressing CD45RA cells (CD27-, CD45RA+)

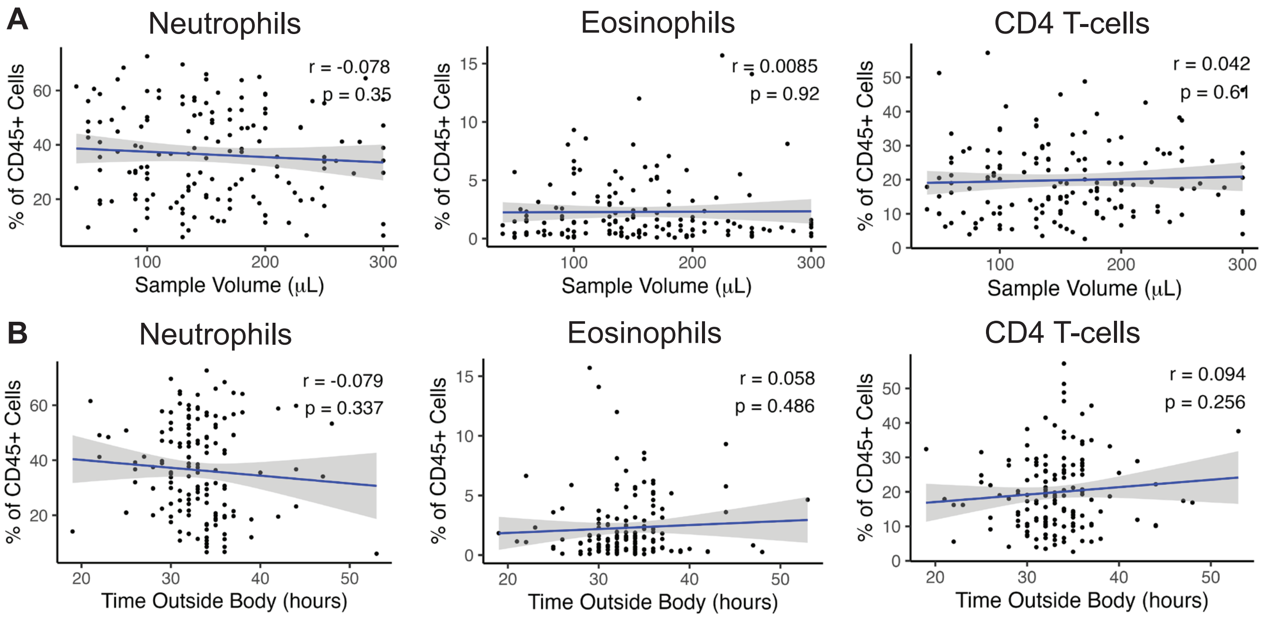


**Supplemental Figure 2: Sample volume and time outside of body does not impact overall cell composition.**

A) Neutrophils, eosinophils, and $CD4^{+}$ T cells proportions in all samples over a range of residual sample volumes.

B) Cells in all samples over the time each sample rested in the clinical lab before processing for this study.

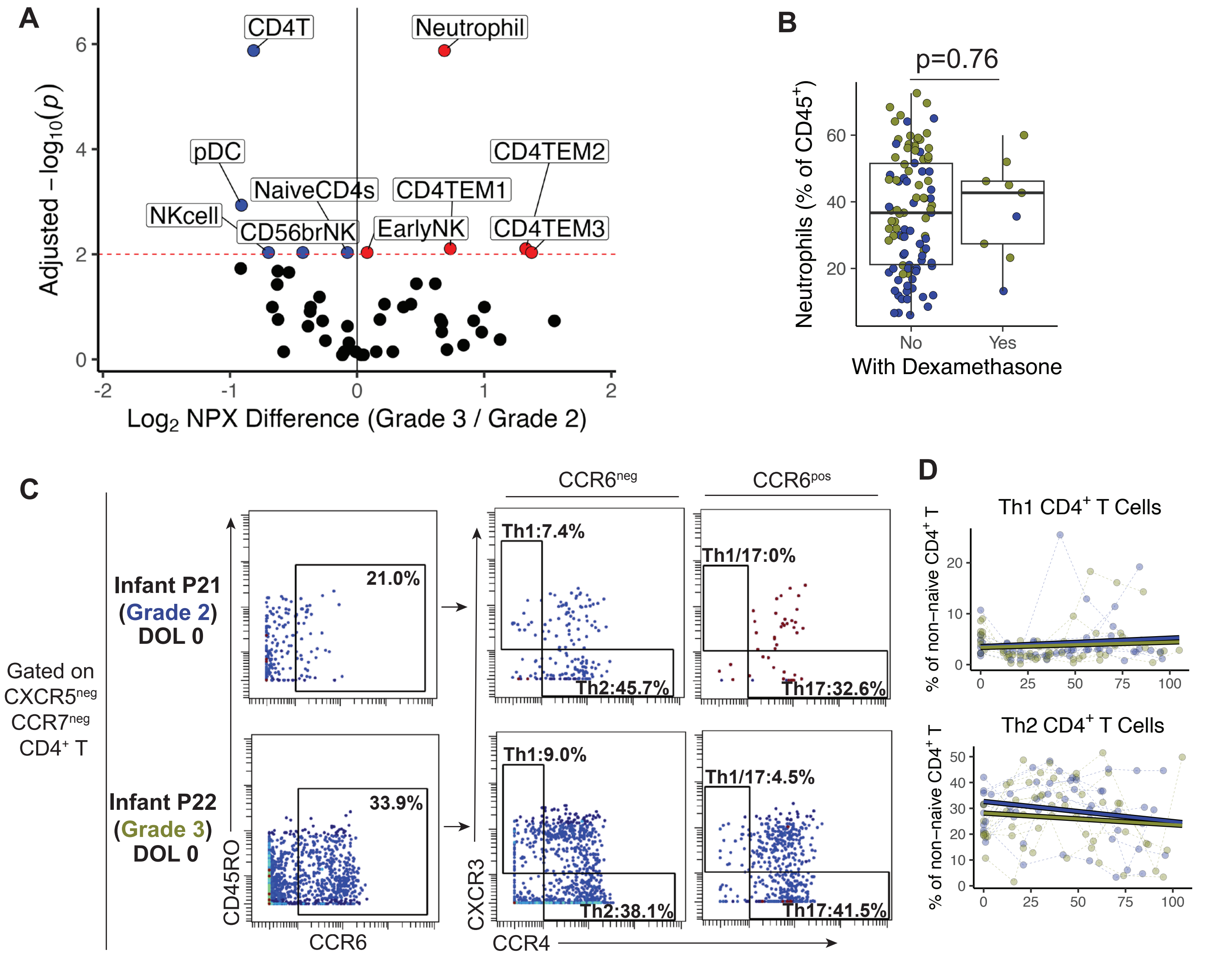


**Supplemental Figure 3: Infants with severe BPD have altered neutrophil and CD4+ T cell profiles**

A) Volcano plot demonstrating cell populations that differ between infants with Grade 3 and Grade 2 BPD. B) Representative CyTOF plots showing the distribution of Th subpopulations within non-naïve CXCR5-, CCR7- T cells from two infants, one ultimately developing moderate BPD (P21) and one severe BPD (P22), at one timepoint on DOL 0. C) Mixed linear model of Th1 and Th2 cells as a proportion of non-naïve CD4+ T cells over time from severe BPD (green) and moderate BPD (blue). D) Proportional neutrophil levels in each sample by whether the sample was collected during active dexamethasone administration. Infants with severe (green) and moderate (blue) BPD shown. P-value derived from Wilcoxon test.
